# Local genetic correlation analysis of Alzheimer’s disease and stroke implicates *PHLPP1* as a shared locus in individuals of African ancestry

**DOI:** 10.64898/2025.12.03.25341552

**Authors:** Nicholas R. Ray, Jiji Kurup, Ajneesh Kumar, Farid Rajabli, Liyong Wang, Wanying Xu, Fulai Jin, Elanur Yilmaz, Caghan Kizil, Luciana Bertholim-Nasciben, Sofia Moura, Aura M. Ramirez, Olusegun Baiyewu, Anthony J. Griswold, Motunrayo Coker, Kyle M. Scott, Kazeem Akinwande, Larry D. Adams, Samuel Diala, Patrice G. Whitehead, Jacob L. McCauley, Mayowa Ogunronbi, Kara L. Hamilton-Nelson, Albertino Damasceno, Andrew F. Zaman, Susan H. Blanton, Albert Akpalu, Michael L. Cuccaro, Kolawole Wahab, Katalina F. McInerney, Karen Nuytemans, Reginald Obiako, Pedro R. Mena, Njideka Okubadejo, Izri M. Martinez, Yared Z. Zewde, Seid A. Gugssa, Fred S. Sarfo, Raj Kalaria, David M. Ndetei, Scott M. Williams, Allison Caban-Holt, ADGC, AfDC, Giuseppe Tosto, Jonathan L. Haines, Rufus O. Akinyemi, Adesola Ogunniyi, Goldie S. Byrd, William S. Bush, Brian W. Kunkle, Jeffery M. Vance, Margaret A. Pericak-Vance, Christiane Reitz

## Abstract

**Background:** Neuropathological studies indicate a strong association between Alzheimer’s disease (AD) and stroke, yet the molecular mechanisms underlying this association remain unclear.

**Methods:** Local genetic correlation analysis was conducted with LAVA using the results from genome-wide association studies on AD and stroke in individuals of African ancestry. Enhanced Hi-C Capture Analysis (eHiCA) examined chromatin interactions using iPSC-derived cells from AD brain autopsy samples.

**Results:** LAVA identified a region shared between AD and stroke on chromosome 18q21.33(*r_g_* = .77, *P* = 2.41×10^-6^). eHiCA demonstrated that the AD and stroke loci interact with regulatory elements in *PHLPP1*. Variants at *PHLPP1* were also associated with AD in an independent set of individuals of African ancestry (*P* = 4.56 × 10^-5^).

**Conclusions:** This study identified a region on top of *PHLPP1* as a locus associated with both AD and stroke. *PHLPP1* inhibits protein kinase B, which contributes to both AD and stroke pathophysiology.

## INTRODUCTION

Alzheimer’s disease (AD) is the leading cause of dementia affecting 6.7 million individuals in the U.S. and incurring an estimated national financial burden of $345 billion annually^1^. AD is a progressive neurodegenerative disorder characterized by pathophysiological changes in the brain (such as the accumulation of amyloid-β plaques and neurofibrillary tangles of hyperphosphorylated tau protein) and cognitive decline (including loss of memory)^2^. Genetic variation plays an important role in AD, with over 75 genetic AD risk loci identified to date^3^. Vascular risk factors and cerebrovascular changes – such as diabetes^4^, hypertension^5^, and stroke^6^ – also predict the onset of AD.

Stroke results in lasting deficits to global cognition, attention, processing speed, memory, language, and motor skills (with the largest rate of decline observed in memory)^7^ and increases the risk of AD by 60%^6^. In addition, cerebrovascular events are associated with more rapid cognitive decline in patients with AD^8^, and postmortem studies indicate that individuals who have both AD pathology as well as cerebral infarcts are more likely to develop dementia than patients with AD pathology alone^9,10^. A genetic correlation (*r_g_*) analysis found a significant global correlation between small vessel stroke and AD (*r_g_* = 0.37) and identified four significant shared pathways between the two traits (three of these involve lipid transport and one involves immune response)^11^. Another study used a gene-based association analysis to identify 16 genes shared between AD and ischemic stroke, many of which act in the immune system^12^.

The exact contribution of cerebrovascular disease to AD remains unknown. AD and stroke share numerous pathophysiological changes in brain tissue, such as hypoperfusion, oxidative stress, immune exhaustion, inflammation, and abnormal expression of amyloid β and tau proteins^13^, which together may lead to altered neurotransmission, synaptic loss, neurodegeneration, and a resulting decline in cognitive ability (i.e., dementia)^14^. It has been hypothesized that by inhibiting the clearance of amyloid, cerebrovascular disease may make an individual more susceptible to AD^15^; and that the two pathologies could act in synergy, increasing the risk of dementia^16^. It is also possible that AD and stroke are two independent, yet convergent, traits with similar pathophysiological processes and shared genetic risk factors^11^.

Ethnicity is another important risk factor for both AD and stroke. Incidence of AD is 64% higher in people of African vs European descent according to a recent meta-analysis^17^. Symptom severity (neuropsychiatric symptoms, cognitive decline, functional impairment) is also greater in African ancestry individuals^18^, who are 50% more likely to experience a stroke and 70% more likely to die from a stroke than European ancestry individuals^19^. Despite this disparity, racial minorities (including individuals of African ancestry) are vastly underrepresented in both dementia^20^ and stroke^21^ research.

To disentangle the mechanistic contribution of CVD to AD in individuals of African ancestry, the current study examined the genetic correlation between stroke and AD in individuals of African ancestry followed by functional fine-mapping of identified shared genetic loci.

## METHODS

### Local Genetic Correlation Analysis of AD and Stroke in African Ancestry

Local genetic correlation between AD and stroke was estimated using LAVA^22^, which uses GWAS summary statistics from multiple traits as input. For AD , summary statistics were obtained for model 1 (controlling for age, sex, and principal components of population stratification) from the most recent and largest GWAS on AD in individuals of African ancestry from the Alzheimer’s Disease Genetics Consortium (ADGC)^23^, which included 2,903 AD cases and 6,265 healthy controls. For stroke, African ancestry-specific GWAS summary statistics were obtained from the GIGASTROKE initiative, a multi-ethnic meta-analysis of all recent major stroke GWAS with a total sample size of 1,614,080^24^, and 3,961 stroke cases and 20,030 healthy controls of African ancestry. Both summary statistics used genome build GRCh37 and a description of the datasets included in both GWAS can be found in their corresponding publications^23,24^. LAVA partitioned the genome into 2,687 semi-independent blocks based on LD using the 1,000 genomes phase 3 African reference panel^25^ and estimated the local genetic correlation between and AD and stroke in each of these regions. The Bonferroni corrected *P*-value threshold for indicating significant genetic covariance of these LD blocks between both traits was derived by dividing .05 by 2,687 or *P* < 1.86×10^-5^.

### Impact of Genetic Ancestry at Regions of Shared Genetic Overlap

The African ancestry AD GWAS used in this study included not only 16 cohorts comprised of African American individuals, but also one cohort from Ibadan, Nigeria^23^. To assess the impact of degree of African ancestry, the pattern of effects in regions of genetic overlap between AD and stroke were compared between the African American cohorts (2,826 cases; 5,637 controls) from the AD GWAS and the Ibadan cohort (77 cases; 628 controls).

Significant regions of overlap resulting from LAVA were also followed up in an independent meta-analysis conducted by the African Dementia Consortium (AfDC; https://africandementiaconsortium.org/). Whole genome sequencing (WGS) data (n = 373) and TOPMED imputed array data from the Recruitment and Retention for Alzheimer’s Disease Diversity Genetic Cohorts in the Alzheimer’s Disease Sequencing Project^26,27^ (n = 180) were collected from 553 Africans from Nigeria, Ghana, Ethiopia, Kenya and Mozambique (275 clinically diagnosed AD cases; 278 controls). The minimum age requirement for this analysis was 55 for AD cases (*M_age_* = 72.9, *SD_age_* = 8.7) and 65 for cognitively normal controls (*M_age_* = 76.7, *SD_age_* = 8.1). Association analyses were run separately for the WGS and imputed datasets using SAIGE and controlling for age, sex, and population stratification as measured by PCs 1-3. Results were then meta-analyzed using Metasoft.

Finally, to examine whether any significant regions of genetic overlap in individuals of African ancestry are also involved in AD or stroke etiology in individuals of European ancestry, results from the largest AD GWAS on individuals of European ancestry (111,326 clinically diagnosed/proxy AD cases; 677,663 healthy controls)^3^ and the European ancestry-specific results from GIGASTROKE GWAS ( 73,652 stroke cases and 1,234,808 healthy controls)^24^ were examined.

### RNA Expression

Genes implicated in the local genetic correlation analysis were first followed up using the Agora AD knowledge portal (https://agora.adknowledgeportal.org/). Agora uses more than 2,100 RNA-seq samples collected from over 1,100 AD cases and controls from three studies: The Religious Orders Study and Memory and Aging and Project^28^, the Mayo RNAseq study^29^, and the Mount Sinai Brain Bank^30^. Agora harmonizes these samples and reports both overall expression and differential expression between AD cases and controls across nine brain regions: Anterior cingulate cortex (ACC), cerebellum (CBE), dorsolateral prefrontal cortex (DLPFC), frontal pole (FP), inferior frontal gyrus (IFG), posterior cingulate cortex (PCC), parahippocampal gyrus (PHG), superior temporal gyrus (STG), and temporal cortex (TCX). Agora also calculates a target AD risk score that ranges from 0-5 and is based on various multi-omic results, including GWAS studies, QTL studies, phenotypic evidence from human and/or animal models, transcriptomic data from RNA-seq profiling, and proteomic data from label-free quantitation (LFQ) and Tandem Mass Tagging (TMT) shot-gun profiling methods^31^.

RNA expression was also analyzed across different cell types using publicly available human single nucleus sequencing data (GSE157827)^32^ and zebrafish single cell sequencing data that have been enriched for gliovascular cells (GSE225721)^33^. *PHLPP1* expression levels were assessed for 9 cell types in humans: astroglia, microglia, oligodendrocytes (OD), oligodendrocyte progenitor cells (OPCs), excitatory neurons, inhibitory neurons, endothelia, fibroblasts, and pericytes; and 12 cell types in zebrafish: astroglia, microglia, OD, OPC, neurons, immature neurons, neuroblasts, immune cells, endothelia, lymph endothelia, pericytes, vascular smooth muscle cells (vascular SMC). The cells that did not fall into one of these 12 categories were analyzed separately and labeled “other”. In humans, comparisons in expression levels for each cell type were observed between AD cases and age-matched controls; and in zebrafish comparisons were evaluated between animals injected with Aβ42 (AD model) versus those injected with phosphate buffered saline (PBS; control group).

To create the Seurat object for human transcriptome analysis, we used the Seurat package (version 5.0.2) in R (version 4.4.0.10)^34^. We filtered out any cells with less than 200 expressed genes, and with genes expressed in less than 3 cells. Following the normalization of the dataset, the FindVariableFeatures() function was utilized with the 2,000 top variable features. Anchors were identified with the *FindIntegrationAnchors* function and integration was performed using the *IntegrateData* function. We used ‘DoubletFinder’^35^ to remove doublets and performed the rest of the analyses on singlets only. The integrated Seurat object included 101,292 cells (55,716 for AD, and 45,576 for Control) with 30,412 features. The data were scaled using all genes, and 30 PCAs (RunPCA) were identified using the *RunPCA* function in the ‘Seurat’ package^34^.

Thirty-six clusters were identified with resolution 1. The main cell types were defined using *AQP4* and *GFAP* for astroglia; *SLC17A7* and *NRGN* for excitatory neurons; *GAD1* and *GAD2* for inhibitory neurons; *PDGFRB*, *MCAM* and *GRM8* for pericytes; *C3* and *DOCK8* for microglia; *PLP1* and *MOBP* for OD; *PDGFRA* and *VCAN* for OPC; and *FLIT1* and *CLDN5* for endothelial cells.

We used the same methods and parameters as described above for creating a Seurat object with the zebrafish dataset. The main cell types were identified as described elsewhere^33,36,37^. Briefly, we used *s100b* and *gfap* for astroglia; *sv2a*, *nrgna*, *grin1a*, *grin1b* for neurons; *pdgfrb* and *kcne4* for pericytes; *cd74a* and *apoc1* for microglia; *mbpa* and *mpz* for OD; *aplnra* for OPC; *myh11a* and *tagln2* for vascular SMC; *lyve1b* for lymph endothelial cells; and *kdrl* for vascular cells. To find the average, aggregate, and percent expression of the genes, we used *AverageExpression*, *AggregateExpression* and *PrctCellExpringGene* functions were used, respectively.

### Enhanced Hi-C Capture Analysis

To examine chromatin interactions at identified regions of genetic overlap, enhanced Hi-C Capture Analysis (eHiCA) was performed on Hi-C data^38^ from iPSC-derived AD-relevant cells and AD brain autopsy (Brodmann Area 9) samples (described elsewhere^39–41^). Autopsy material was obtained from the Alzheimer’s Disease Research Centers at Emory University, Northwestern University, and the John P. Hussman Institute for Human Genomics. All samples were acquired with informed consent for research use and approved by the institutional review board of each center. In situ Hi-C library was prepared using a protocol adapted from Rao et al with a 4-cutter enzyme^42^. For each library, 340∼860 million paired-end reads at 150 bp length were obtained.

Chromatin loops were called using *HiCorr*^43^ to correct bias and *LoopEnhance*^44^ to remove noise. In addition, layered H3K27ac ChIP-seq data from 7 cell lines from the ENCODE project^45^, H3K27ac and H3K4me3 ChIP-seq data from adult brains with NCBI GEO accession number GSE116825^43^ were used to annotate functional DNA elements near the chromatin loops. Hi-C libraries from eight donors (4 African American and 4 non-Hispanic White) were used for the analysis. Like virtual 4C analysis, eHiCA visualizes chromatin interacting signals from any bait-of-interest except that the signals are extracted from *DeepLoop*-enhanced Hi-C contact matrices. As a result, the plotted signals are enhanced with observed *versus* expected ratios. We typically select a 5kb bin as the bait of interest and show the interacting signal within +/- 2 Mb surrounding the bait. The loop strength within the +/- 2 Mb surrounding regions is visualized by gradient heatmap.

## RESULTS

### Local Genetic Correlation Analysis of AD and Stroke in African Ancestry

To examine the genetic overlap between AD and stroke in individuals of African ancestry, an analysis of local genetic correlation was conducted using LAVA, which identified a single region shared between AD and stroke meeting the Bonferroni corrected *P*-value threshold for multiple testing at chr18:59695907- 61178035 (chr18q21, *r_g_* = .77, *P* = 2.41×10^-6^). Examination of the LD structure and genetic association patterns at this locus identified disease-associated loci exerting an effect in the same direction in both traits. The top signal in the African ancestry AD GWAS^23^ is chr18:60662938 (rs512453, effect allele = T, other allele = C, effect allele frequency = .90, β = 0.25, *P* = 6.02×10^-5^), which is located on the 3’ end of *PHLPP1* and shows strong support from nearby SNPs (**Figure 1A**). According to 1000 Genomes reference data, this variant is common in both Africans and Europeans, but is much less polymorphic in Africans (African allele frequency = .97, European allele frequency = .56)^25,46^. The top signal in the African ancestry stroke GWAS^24^ is chr18:60380912 (rs73465039, effect allele = T, other allele = G, effect allele frequency = .06, β = 0.22, *P* = 1.80×10^-4^), which shows strong support from nearby SNPs in the promoter region of *PHLPP1* (**Figure 1B**). The effect allele for this variant occurs somewhat rarely in Africans (allele frequency = .07) and not at all in Europeans^25,46^. The top SNPs from the African ancestry AD and stroke GWAS are not in LD with each other based on r^2^ and therefore may represent two LD blocks that each show strong regional support by other variants (D′ is high between the top hits from both GWAS however; **Supplementary Figure 1**). Notably, rs80108507, a significant SNP from the African ancestry stroke GWAS that is in high LD with the stroke top hit, is an eQTL affecting *PHLPP1* expression levels in oligodendrocytes^47^. This region of genetic overlap on *PHLPP1* represents a potentially novel AD-risk locus that may inform future research on the shared causal relationship between cerebrovascular health, neurodegenerative disease, and genetic ancestry.

**Figure 1.**
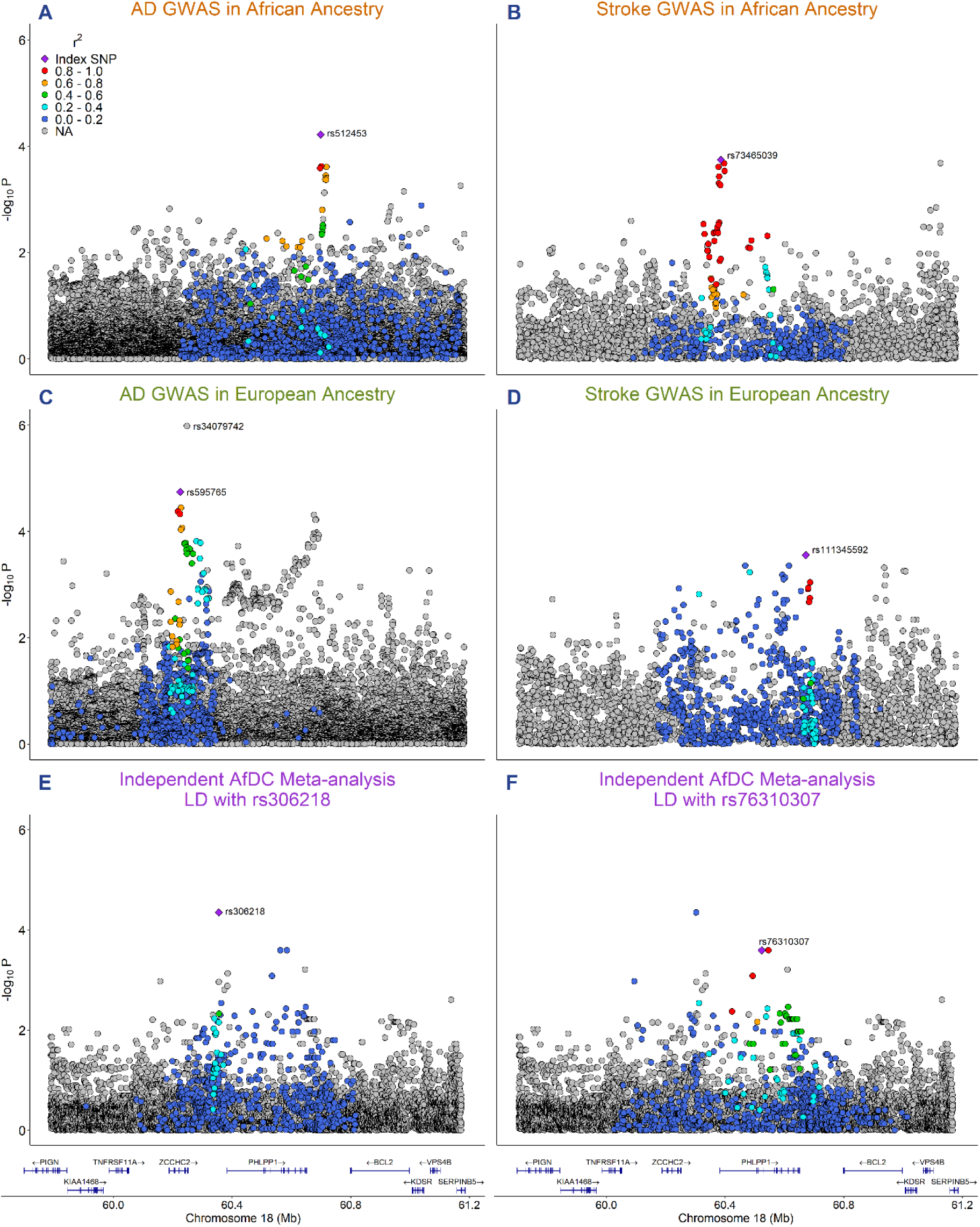
Regional Association Plots at the chr18q21 locus identified in genetic covariance analyses. The -log10 transformed *P*-value is displayed on the y-axis and base-pair position on chromosome 18 is shown on the x-axis using genomic build GRCh37. The color indicates LD in relation to the labeled top hits from each GWAS in the region. Higher r^2^ values are indicated by warmer colors (reds) and lower r^2^ values are indicated by cooler values (blues). **A**) Results from the AD GWAS in African ancestry^23^ that was used in the local genetic correlation analysis. The most significant variant (rs512453; *P* = 6.02×10^-5^) shows strong support from nearby SNPs on the 3’ end of *PHLPP1*. **B**) Results from stroke GWAS in African ancestry^24^ that was used in the local genetic correlation analysis. The most significant variant (rs73465039; *P* = 1.80×10^-4^) shows strong support from nearby SNPS in the promoter region of *PHLPP1*. **C**) Results from the AD GWAS in Europeans^3^. The most significant variant (rs34079742, *P* = 1.04×10^-6^) is not present in 1000 Genomes, so this plot shows LD in relation to the second most significant variant (rs595765; *P* = 1.80×10^-5^). **D**) Results from the stroke GWAS in Europeans^24^. **E**) Results from an independent meta-analysis of 553 Africans performed by the AfDC. This plot shows LD associated with rs306218 (*P* = 4.46×10^-5^), which is located between *ZCCHC2* and *PHLPP1*. **F**) Same results as in E, but this plot shows LD associated with rs76310307 (*P* = 2.55×10^-4^), which is located on top of *PHLPP1* and shows strong regional support from nearby SNPs.

### Impact of African Ancestry in AD

Because the AD GWAS used in the local genetic correlation analysis contains a cohort from Ibadan, Nigeria^23^, we were able to leverage those results to examine the impact of degree of African ancestry, which is known to be greater in continental Africans compared to African Americans^48^. **Supplementary Figure 2A** shows the regional association plot from the AD GWAS at the *PHLPP1* locus for the African American cohorts sampled in the U.S. while **Supplementary Figure 2B** shows the local association results for the African (Ibadan) cohort with higher degree of African ancestry. **Supplementary Figure 2C** shows the beta coefficients and standard errors for the top SNP from the US cohorts (rs512453, effect allele = T, other allele = C, effect allele frequency = .90, β = 0.25, SE = 0.06, *P* = 5.01×10^-5^) across all cohorts from the African GWAS in which this SNP was present (Ibadan included), and **Supplementary Figure 2D** shows the beta coefficients and standard errors for the top SNP from the Ibadan cohort (rs183411323, effect allele = C, other allele = G, effect allele frequency = .004, β = 4.23, SE = 1.17, *P* = 3.13×10^-4^). These analyses show both different top variants and patterns of effects between the US and Ibadan cohorts, suggesting that degree of African ancestry may be modulating the effect in this region^48^. The sample size of the Ibadan cohort (N = 705) is small compared to the US cohorts (N = 8,463) however, and so this result should be interpreted with caution. Reference data shows that the top hit from the US cohorts (rs512453) is common in both Africans (allele frequency = .97) and Europeans (allele frequency = .56), while the top hit from the Ibadan cohort (rs183411323) is only present in Nigerians (allele frequency in Esan = .01; allele frequency in Yoruba = .009)^25,46^.

### Genetic Association at Chr18q21 in the AfDC Meta-Analysis

To further assess the impact of African ancestry in AD and follow up on the results from the local genetic correlation analysis, the chr18q21 region was examined using results from an independent meta-analysis of 553 Africans conducted by the AfDC. Visualization of these results reveal two potentially distinct LD blocks: one located between *ZCCHC2* and *PHLPP1* the with top variant chr18:60297877 (rs306218, effect allele = T, other allele = C, effect allele frequency = .18, β = 0.75, *P* = 4.46×10^-5^; **Figure 1E**), and another located in *PHLPP1* with top variant chr18:60519473 (rs76310307, effect allele = C, other allele = G, effect allele frequency = .06, β = 1.03, *P* = 2.55×10^-4^; **Figure 1F**). The chr18q21 region results from this analysis are reported in **Supplementary Table 1** along with the results from the AD^3,23^ and stroke GWAS^24^, as well as the Ibadan cohort from the AD GWAS in African ancestry^23^. Significant variants resulting from the AfDC meta-analysis are not overlapping with the significant variants from the African ancestry AD^23^ and stroke^24^ GWAS. Some of these variants are missing in the other datasets, but most are not significant and not showing LD with the top hits from the African ancestry AD and stroke GWAS according to r^2^ estimates based on 1000 Genomes reference data^25^. As the AD and stroke GWAS were conducted on mostly African American participants, and the AfDC meta-analysis was conducted on participants from five African countries, one reason we may not observe overlap could be that the degree of African ancestry may be modulating the effect in this region. However, it should be noted that we also do not observe overlap between significant results from the AfDC meta-analysis and significant results from the Ibadan cohort of the AD GWAS – one potential explanation for this could be that the 3 most significant variants from the Ibadan cohort of the AD GWAS were filtered out from the AfDC meta-analysis results during the quality control pipeline (**Supplementary Table 1**).

### Association of Genetic Variation at Chr18q21 with AD and Stroke in Individuals of European Ancestry

To determine whether there is also a relationship between AD and stroke in individuals of European ancestry at chr18q21, results from European AD^3^ and stroke^24^ GWAS were also examined in this region. The top hit in the European stroke GWAS^24^ in the chr18q21 region is chr18:60668973 (rs111345592, AF = .75, β = -0.03, SE = 0.01, *P* = 2.78×10^-4^) with strong regional support with a group of other SNPs near the 3’ end of *PHLPP1* (**Figure 1D**). The top hit in the European AD GWAS^3^ at chr18q21 is chr18:60184001 (rs34079742, AF = .36, β = -0.05, *P* = 1.04×10^-6^; **Figure 1C**). This SNP is not present in 1000 Genomes, but examination of other top SNPs reveals three distinct LD blocks in this region: One on top of *ZCCHC2* (rs595765, allele frequency = .62, β = 0.04, *P* = 1.80×10^-5^, **Supplementary Figure 3A**), one on the 5’ end of *PHLPP1* (rs144225393, allele frequency = .06, β = -0.06, *P* = 1.88×10^-4^, **Supplementary Figure 3B**), and one on the 3’ end of *PHLPP1* (rs72343350, allele frequency = .08, β = -0.06, *P* = 4.90×10^-5^, **Supplementary Figure 3C**). The results of both the European and African ancestry AD and stroke GWAS are reported in **Supplementary Table 1** along with the results from the independent AfDC meta-analysis. Many significant SNPs (*P* < 5×10^-4^) from the European ancestry AD GWAS are also nominally significant in the European ancestry stroke GWAS (rs595765, rs1108168, rs9951519, rs4941139, rs1053951, rs12150719, rs7226431, rs2051468, rs7227643, rs4941141, rs1606890, rs1473980, rs9944724; see **Supplementary Table 1**), indicating that some genetic overlap between the two traits also exists in individuals of European ancestry at chr18q21. Although both ancestries show disease association within *PHLPP1*, the variants most strongly associating with either AD or stroke differ between the European ancestry GWAS and the African ancestry GWAS (see **Supplementary Table 1**).This suggests that different genetic variants may be affecting the relationship between AD and stroke in individuals of European ancestry compared to those in individuals of African ancestry.

### RNA Expression of *PHLPP1* in Humans and Zebrafish

The top hits from the African ancestry AD^23^ and stroke^24^ GWAS in the region resulting from the local genetic correlation analysis were both in *PHLPP1*, which is highly expressed across nine brain regions (**Supplementary Figure 4A**). *PHLPP1* is differentially expressed between AD cases and controls in seven of these regions (**Supplementary Figure 4B)** including the DLPFC (*P* = 2.99 × 10^-6^) and the PHG (*P* = 2.07 × 10^-11^), which are commonly affected by AD. In line with this notion, the target AD risk score for *PHLPP1,* indicating its likelihood of being involved in the etiology of AD, is relatively high at 3.27 out of 5 (https://agora.adknowledgeportal.org/).

To examine expression of *PHLPP1* in AD and stroke relevant cell types, single cell RNA sequencing data was examined in both humans and zebrafish, which are widely used in genetic studies of human disease as 82% of disease-related genes in humans have a zebrafish ortholog^49^. *PHLPP1* is highly expressed across multiple cell types relevant to stroke and AD in humans, including neurons, OD, and astroglia (**Supplementary Table 2** and **Supplementary Figure 5)**. Significant expression change between AD cases and controls is observed in OD (*P* = .01; **Supplementary Figure 6**), where expression is upregulated in controls compared to AD cases. In zebrafish, *phlpp1* is also highly expressed in astroglia (**Supplementary Table 3** and **Supplementary Figure 7)** where it is expressed significantly higher (*P* = .002; **Supplementary Figure 8**) in the AD model compared to the control model. Together these results suggest that *PHLPP1* is highly expressed in brain tissues and cell types that are relevant to both AD and stroke pathophysiology.

### Enhanced Hi-C Capture Analysis at Chr18q21

Results of the eHiCA analysis are shown in **Figure 2**. The bait used for the stroke top hit is in the promoter region of *PHLPP1* and shows strong chromatin interaction with the 3’ end of the gene. The reverse is true for the bait used for the AD top hit, which is located at the 3’ end of *PHLPP1* and interacts strongly with the promoter region. Not only do both baits interact strongly with each other, but they also both interact with the same chromosomal regions of six additional genes: the promoter region of *PIGN*, *RELCH*, *BCL2*, *VPS4B*; and the 3’ end of *ZCCHC2* and *KDSR*. A similar pattern of chromatin interactions, indicating a coordinated regulation of the same set of genes, was observed in brains across ancestries (African and European), and in different AD-relevant cell types (microglia, neurons, and OD) consistent with the RNA expression data from human brain described above.

**Figure 2.**
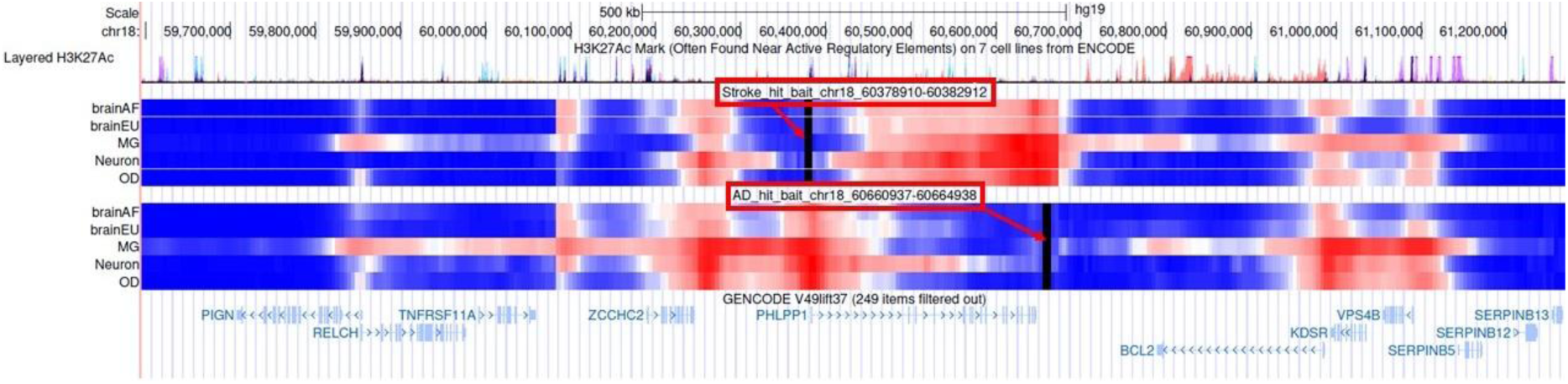
Chromatin interactions for the top SNPs from the AD and stroke GWAS in the region showing functional overlap between the two traits. The baits for the top SNPs are indicated by the two solid vertical black lines, which are further highlighted by the red arrows. The bait for the AD top SNP (rs512453) is shown on the top of the figure while the bait for the stroke top SNP (rs73465039) is shown on the bottom. Chromatin interactions with each bait are displayed by the horizontal bars, where warm colors (e.g., red) indicate stronger interaction and cooler colors (e.g., blue) indicate weaker interaction. Chromatin interactions are shown for two ancestries – African (brainAF) and European (brainEU) – and three AD-relevant cell types – microglia (MG), neurons, and oligodendrocytes (OD).

## DISCUSSION

Local genetic correlation analysis identified a region of genetic overlap between AD and stroke in individuals of African ancestry on chromosome 18q21. While the stroke top hit is in the promoter of *PHLPP1* and the AD top hit is on the 3’ end of the gene, eHiCA analysis shows strong chromatin interactions not only between the two top hits, but also with functional elements in six surrounding genes. This suggests that both the AD and stroke loci are functionally modifying the same set of genes at and around *PHLPP1*.

*PHLPP1* belongs to the Pleckstrin Homology domain Leucine-rich repeat Protein Phosphatases (PHLPP) family, which inhibit protein kinase B (AKT)^50^, protein kinase C (PKC)^51^, and MAPK^52^. AKT plays a role in cell survival and growth signaling, and its dysregulation is associated with the development and progression of multiple neurodegenerative diseases, including AD^53,54^. AKT protects neurons from Aβ-induced neurotoxicity, and activation of the AKT pathway is thought to delay the progression of AD^54,55^. AKT also promotes angiogenesis and can potentially relieve oxidative stress associated with stroke^56,57^. Deletion of *PHLPP1* in mice resulted in enhanced AKT activation and reduced neurovascular damage after middle cerebral artery occlusion^58^. In line with these notions, *PHLPP1* is highly expressed across brain regions, is highly expressed in AD and stroke relevant cell types, and is differentially expressed between AD cases and controls.

Analyses contrasting the local association patterns at the AD locus in African Americans with individuals with higher degree of African ancestry showed different patterns of effects, suggesting that local and/or global ancestry may be modulating the effect at this locus. Our independent meta-analysis of WGS data from Africans identified an LD block on top of *PHLPP1* that did not overlap with the African ancestry AD GWAS that was conducted mostly in African Americans^23^. Examination of this region in the largest available GWASs in individuals of European ancestry revealed suggestive signals at *PHLPP1* in both AD^3^ and stroke^24^, but with a different set of variants than those shown in the African ancestry GWAS. We also identified this locus as one of two genome-wide significant loci in analyses using admixture mapping to prioritize genetic regions associated with AD in African Americans^59^. This evidence points to an effect of genetic ancestry on the relationship between AD and stroke at chr 18q21 and highlights the importance of examining genetic relationships across a diverse range of ancestral populations.

Analysis of chromatin interactions with the top hits from the African ancestry AD and stroke GWAS indicate that the two regions carrying the disease associated haplotypes interact with each other in the *PHLPP1* gene: the stroke top hit shows strong chromatin interaction with the genomic region in the 3’ end of the gene where the AD top is located, and the AD top hit shows strong chromatin interaction with the genomic region in the 5’ end of the gene where the stroke top hit is located (**Figure 2**). Both top hits are also interacting with the same functional elements of six nearby genes: the promoter region of *PIGN*, *RELCH*, *BCL2*, *VPS4B*; and the 3’ end of *ZCCHC2* and *KDSR*. *PIGN* encodes a protein that transports phosphatidylethanolamine (PE), which is elevated in individuals with *PSEN1* and *PSEN2* mutations, suggesting that PE may increase γ-secretase activity and Aβ production^60,61^. *RELCH* is involved in cholesterol transport^62,63^, a causative pathway involved in both AD^64^ and stroke^65^ etiology. *ZCCHC2* encodes a protein that belongs to the zinc finger CCHC-type (ZCCHC) superfamily, which regulates zinc and RNA metabolism^66^. Zinc metabolism is important for neurogenesis, neurotransmission, and memory formation^67^; and a GWAS from the Mexican Health and Aging Study found a strong association between episodic memory and *ZCCHC2*^68^. *BCL2* encodes a mitochondrial protein that promotes cell survival by regulating apoptosis^69^ and blocking BCL2 reduced tau burden in a mouse model of AD^70^. *KDSR* encodes 3-Ketodihydrosphingosine reductase, an enzyme involved in the synthesis of sphingolipids, which are abundant in the central nervous system and play an important role in a variety of cellular functions, such as membrane trafficking, apoptosis, and cell differentiation and proliferation^71^. Dysfunction of sphingolipid metabolism results in neurodegeneration^72^, with high levels of ceramides, for example, leading to memory loss^73^, decreased hippocampal volume, cognitive decline^74^, and increased risk of AD^75^. *VPS4B* encodes a member of the ATPases Associated with diverse cellular Activities (AAA) protein family; is part of the endosomal sorting complexes required for transport (ESCRT) system; and is involved in a number of important cellular functions, such as multivesicular body formation, lysosomal degradation, and intracellular protein trafficking^76–79^. Reduced *VPS4B* expression may cause impaired endosomal sorting, neuroinflammation, and necroptosis^80^ leading to neurodegeneration that can contribute to AD^81,82^. Alternatively, increased *VPS4B* expression was observed in the hippocampus after cerebral artery occlusion in adult rats, and knockdown of *VPS4B* resulted in down-regulation of active caspase-3 (an enzyme involved in apoptosis) expression, suggesting that upregulation of this gene may contribute to oxygen-glucose deprivation-induced cell death in stroke^83^.

A single gene can influence multiple traits, a phenomenon known as pleiotropy that is ubiquitous in the study of human genetics and disease^84^. It is important to study pleiotropy to understand the relationship between multiple phenotypes and potentially uncover biological pathways that are shared between them. Past studies have examined genetic overlap between AD and ischemic stroke, but these studies were mostly conducted on individuals of European ancestry and estimated global correlation. Global correlation examines the relationship between two traits across the entire genome but does not consider local relationships at specific genetic loci. The present study utilized genome-wide LD information to partition the genome into semi-independent LD blocks and then estimated the correlation between AD and stroke at each of these regions, allowing the discovery of potential local relationships that may be missed by averaging effects across the entire genome. As the partitions are ∼1 MB on average, the possibility remains that we missed additional correlations between smaller loci that are diluted by including additional variants in the 1MB window.

The current study nominates *PHLPP1* as a potential therapeutic target shared between AD and stroke in both individuals of African and European ancestry. Our analyses suggest that the disease-associated genetic variation at this locus is modified by genetic ancestry, and in individuals of African ancestry also by degree or origin of African ancestry. It is possible that by inhibiting AKT, *PHLPP1* is acting in a causal pathway for both AD and stroke. Evidence suggests that suppression of this gene increases AKT activation, potentially leading to more favorable outcomes in both diseases. eHiCA analysis suggests that the genomic regions underlying the top hits in *PHLPP1* are also interacting with functional elements in nearby genes. Further research is warranted to determine how these genes contribute to a shared mechanistic pathway between AD and stroke, how genetic ancestry effects this mechanism, and whether this locus should be considered as a target for future interventions.

## Data Availability

This study used only publicly available data. The summary statistics from the African ancestry stroke GWAS are available in the GWAS Catalog (https://www.ebi.ac.uk/gwas/studies/GCST90104549)and the summary statistics from the African AD GWAS are available through the National Alzheimer's Coordinating Center (naccdata.org).

https://www.ebi.ac.uk/gwas/studies/GCST90104549

https://naccdata.org/

## ACKNOWLEDGEMENTS

The National Institute on Aging supported this work through the following grants: U01AG084545, U19AG074865, R01AG072547, U01AG072579, RF1AG059018, AG072959, AG058654, U01AG052410, AG057659.

This work was funded in part by the Hussman Foundation.

The results published here are in whole or in part based on data obtained from Agora, a platform initially developed by the NIA-funded AMP-AD consortium that shares evidence in support of AD target discovery. Agora is available at: doi:10.57718/agora-adknowledgeportal.

The authors have no conflicts of interest to report related to this publication.

## Notes

### Competing Interest Statement

The authors have declared no competing interest.

