## Supplemental FIgures for "Local genetic correlation analysis of Alzheimer’s disease and stroke implicates *PHLPP1* as a shared locus in individuals of African ancestry"

### **Supplementary Figure 1.** LD Matrix of Top Hits from the African Ancestry AD and Stroke GWAS at Chr18q21.

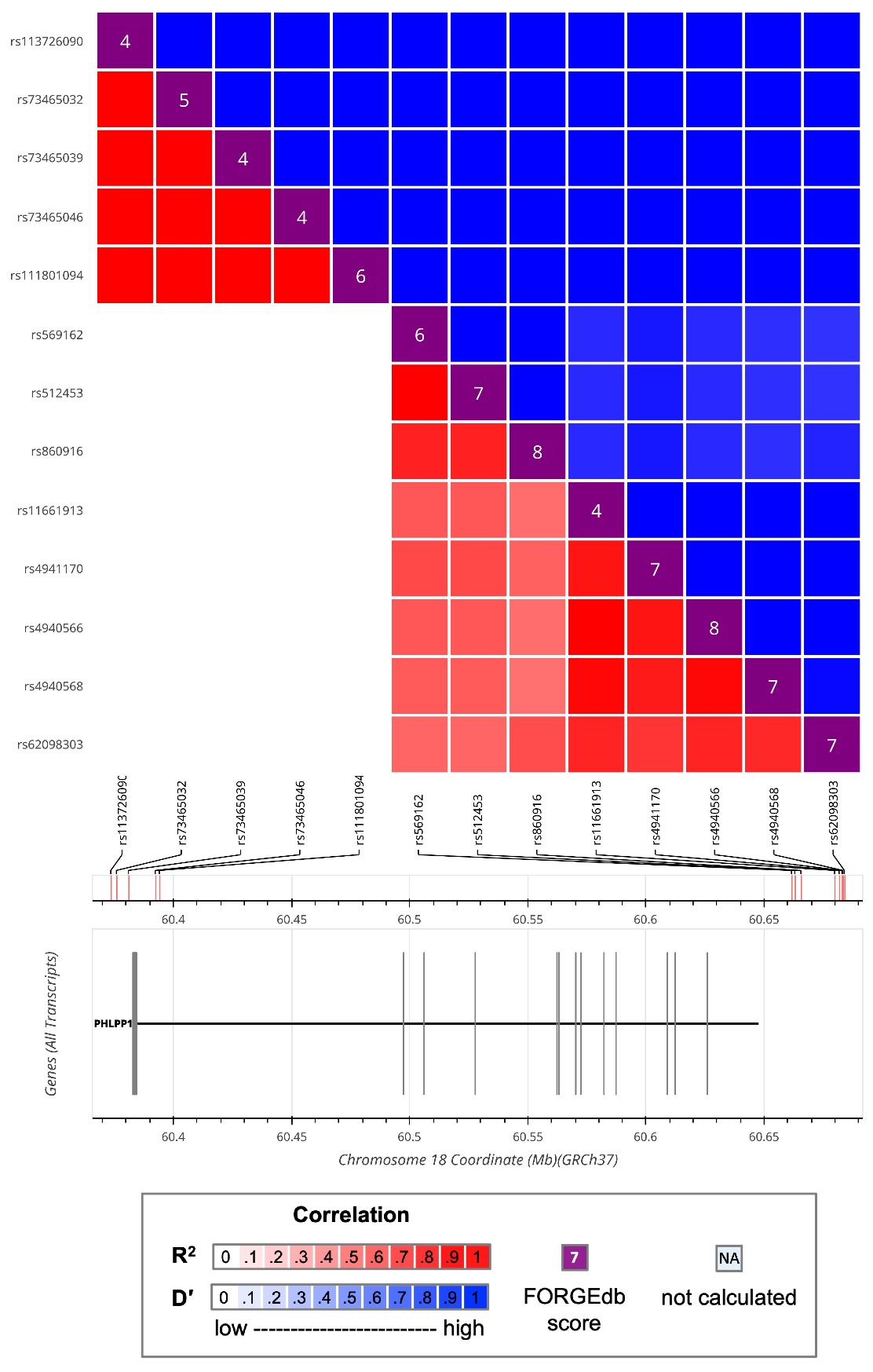

This plot was generated using LDlink (https://ldlink.nih.gov) employing the LDmatrix tool and selecting the 1000 Genomes^1^ African populations as reference. Top hits were defined using a *P*-value threshold of *P* < 5×10^-4^. AD top hits: rs512453, rs860916, rs62098303, rs569162, rs4941170, rs4940568, rs11661913, rs4940566. Stroke top hits: rs73465039, rs73465046, rs113726090, rs111801094, rs73465032. Stroke top hit rs150183720 was omitted from this figure as it was not in proximity nor was it in LD with the rest of the SNPs shown here. The pattern of LD suggests two distinct haplotypes between the stroke top hits and the AD top hits.

### **Supplementary Figure 2.** Comparison of Effects at Chr18q21 Between African American Cohorts in the US and Africans from Ibadan, Nigeria.

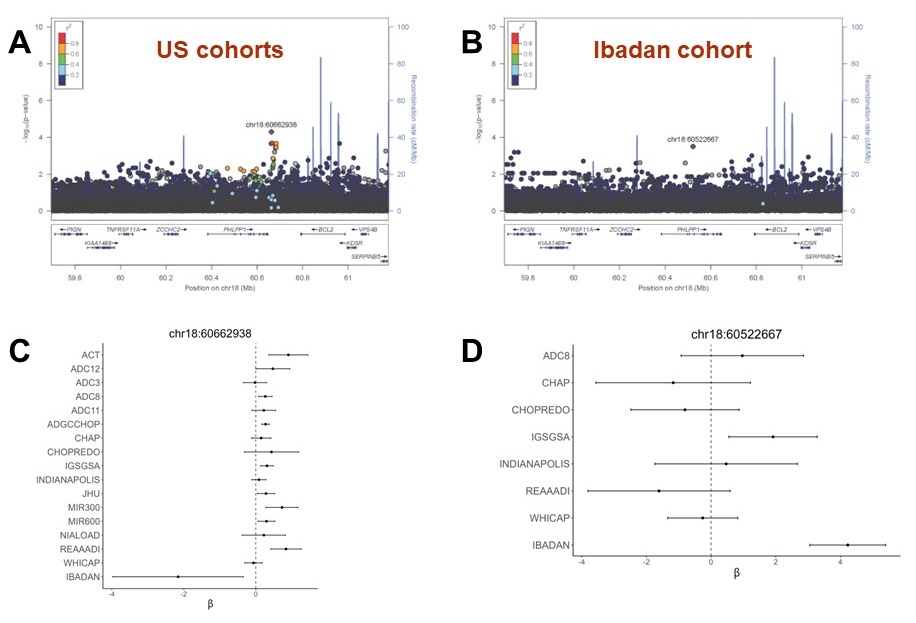

A) Regional association plot for chr18:59695907-61178035 for all US cohorts from the African ancestry ADGC GWAS^2^. B) Regional association plot for the cohort of Africans from Ibadan, Nigeria. C) Betas and standard errors for the top SNP in this region in US cohorts across all cohorts from the African AD GWAS in which this SNP was present. D) Betas and standard errors for the top SNP in this region in the Ibadan cohort across all cohorts from the African AD GWAS in which this SNP was present.

### **Supplementary Figure 3.** Regional Association Plots for AD in Europeans at Chr18q21.

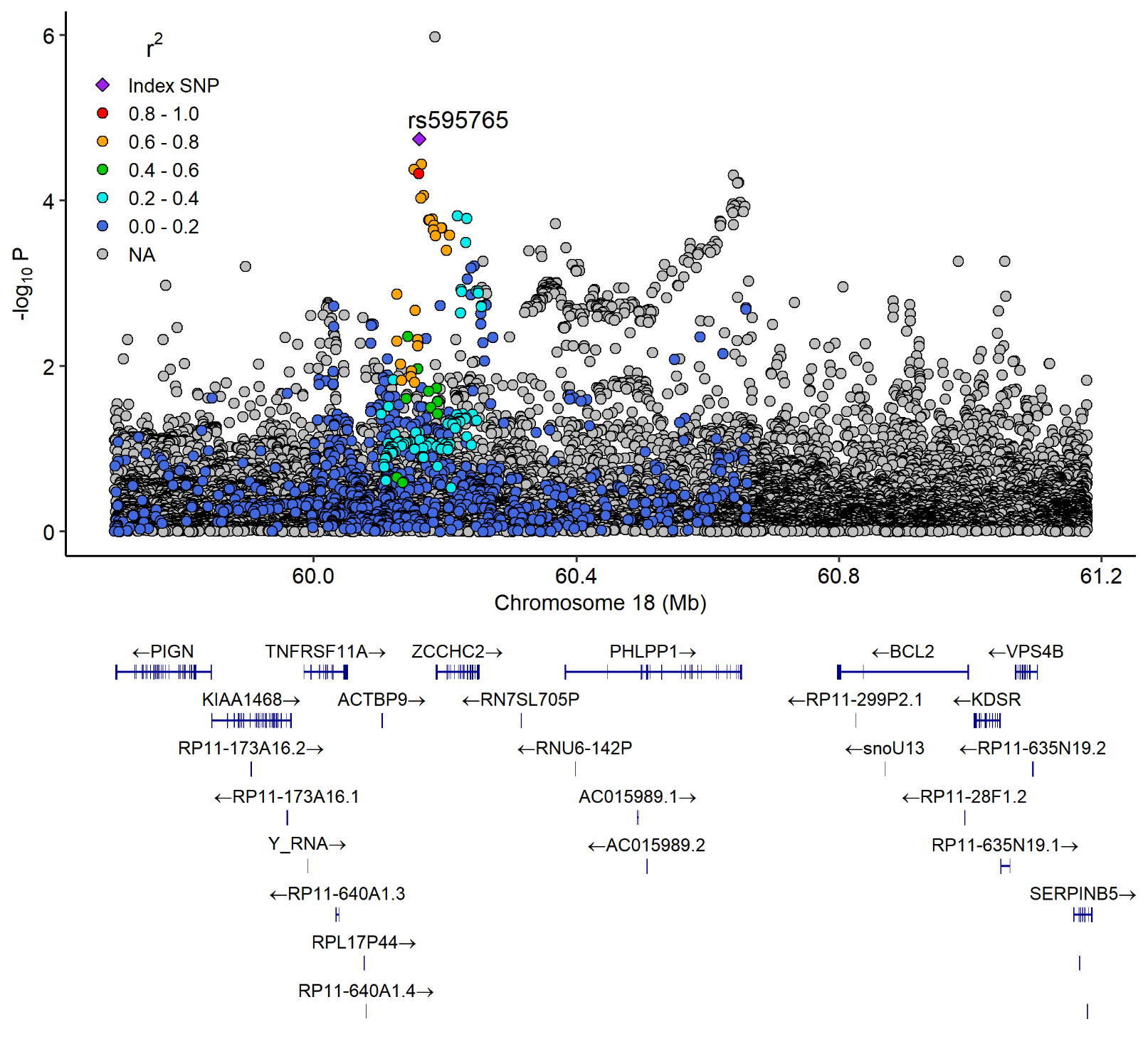

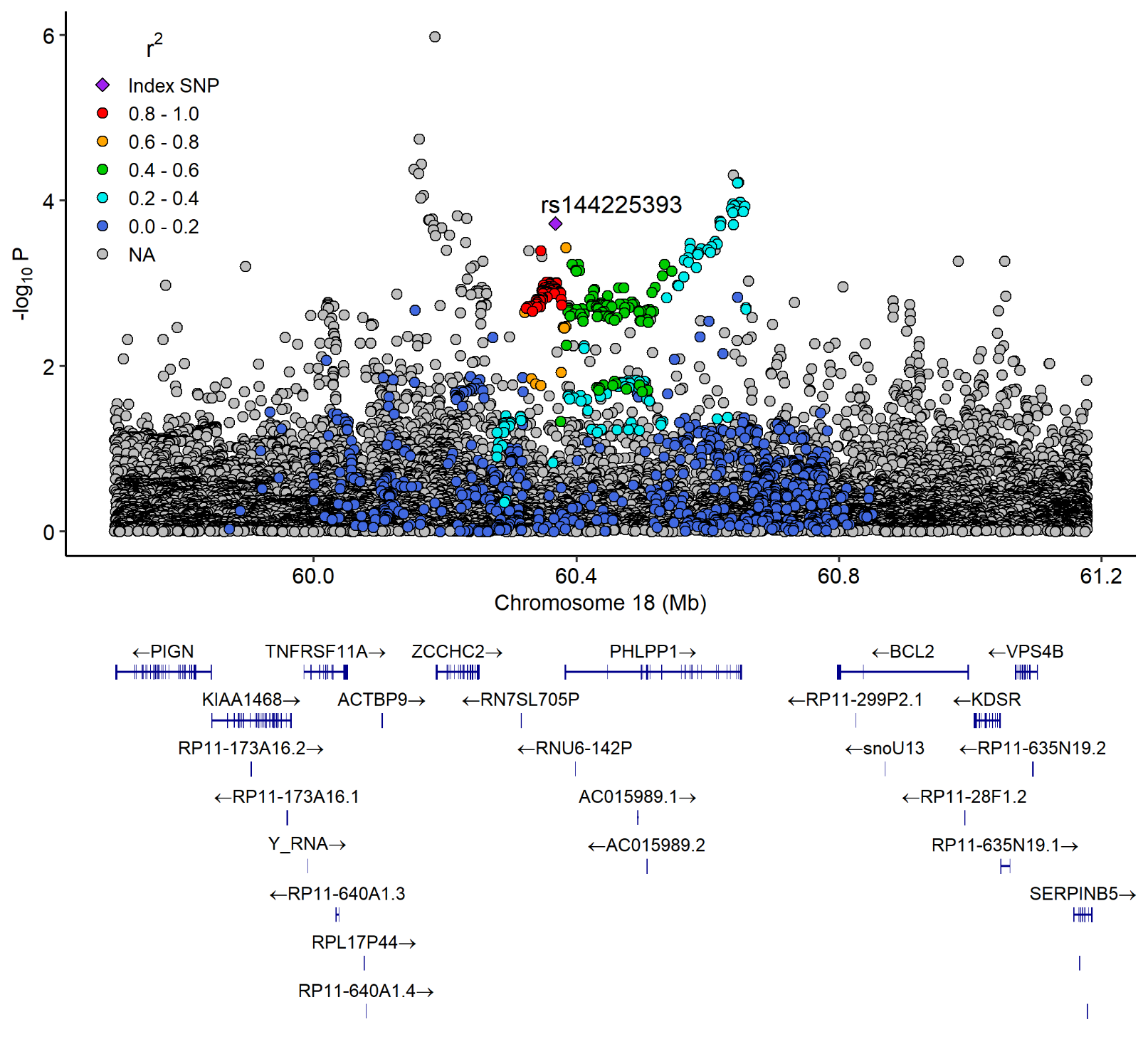

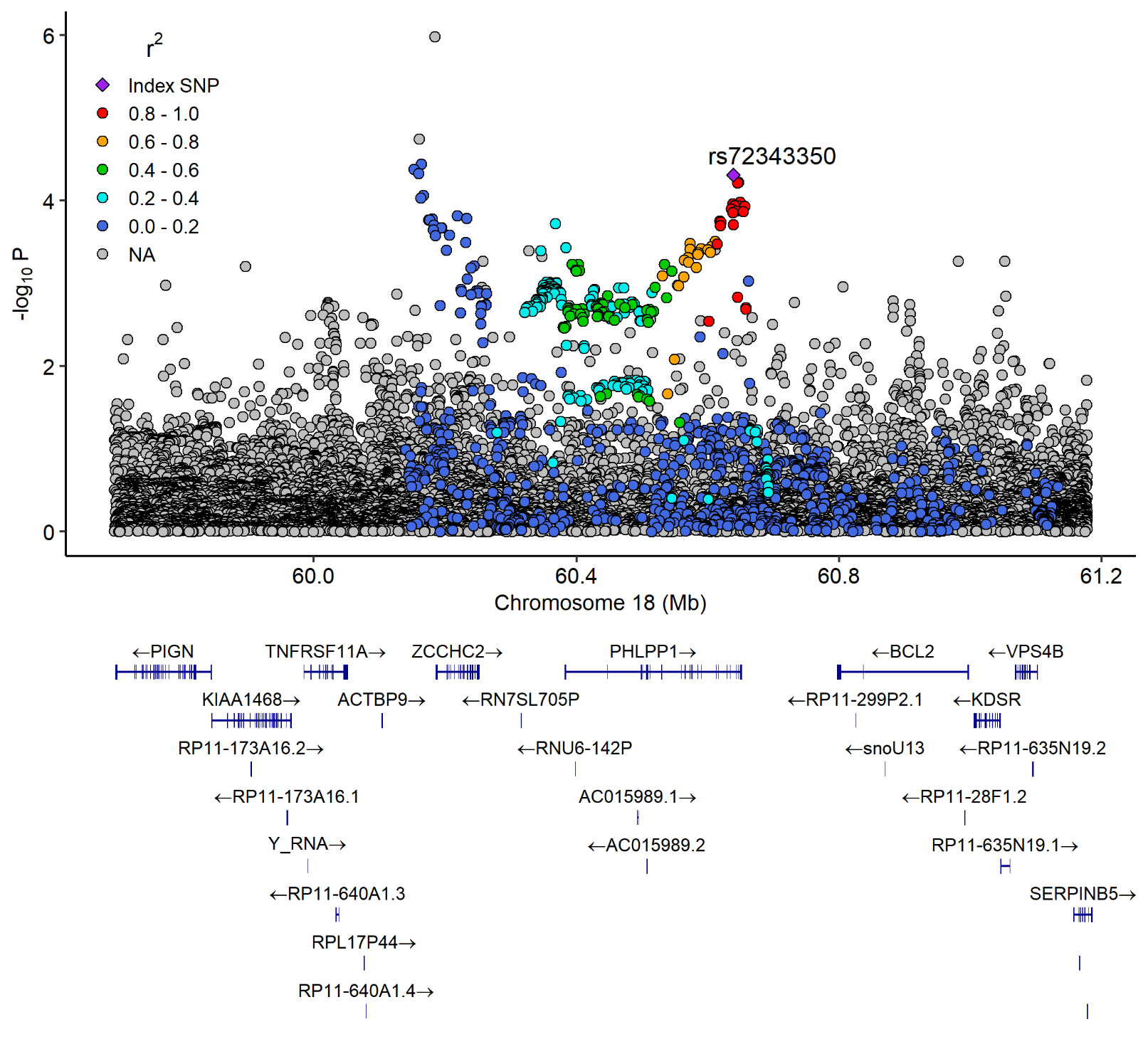

Regional association plots showing three haplotypes from the European AD GWAS results^3^ at chr18q21. **A** shows LD patterns in relation to rs595765 (*P* = 1.80×10^-5^) on top of *ZCCHC2*; **B** shows LD patterns in relation to rs144225393 (*P* = 1.88×10^-4^) on the 5’ end of *PHLPP1*; and **C** shows LD patterns in relation to rs72343350 (*P* = 4.90×10^-5^) on the 3’ end of *PHLPP1*. The -log10 transformed *P*-value is displayed on the y-axis and base-pair position on chromosome 18 is shown on the x-axis. The color indicates LD in relation to the labeled top hits, with higher r^2^ values indicated by warmer colors and lower r^2^ values indicated by cooler values.

### **Supplementary Figure 4.** RNA Expression of *PHLPP1* Based on Agora Evidence

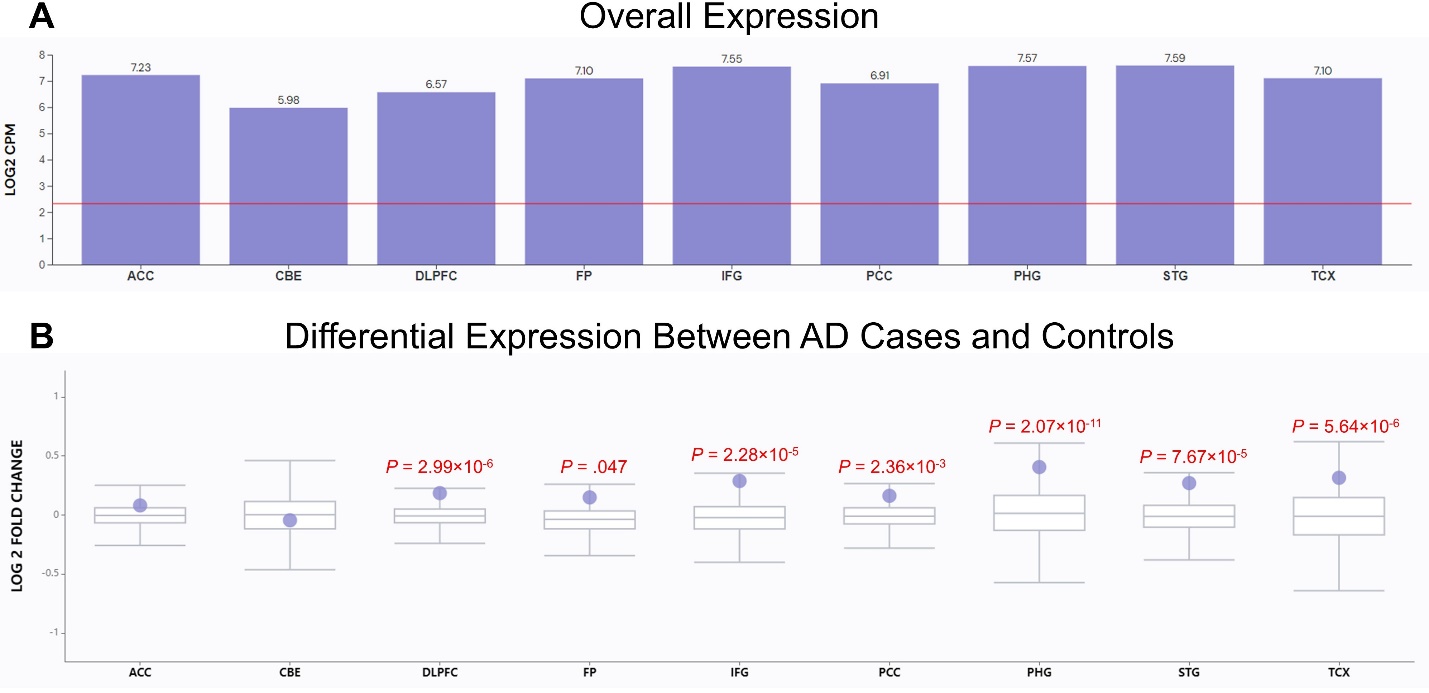

A) Median RNA-seq read counts per million (CPM) reads for *PHLPP1* across 9 brain regions. Meaningful expression is defined as a log2 CPM greater than log2(5), which is shown by the red line. B) Log 2 Fold Change showing the differential expression of *PHLPP1* between AD cases and controls in each brain region. *P*-values are shown in red for regions with a significant Log 2 Fold Change value.

### **Supplementary Figure 5.** Overall RNA Expression of *PHLPP1* Across 9 Cell Types in Humans

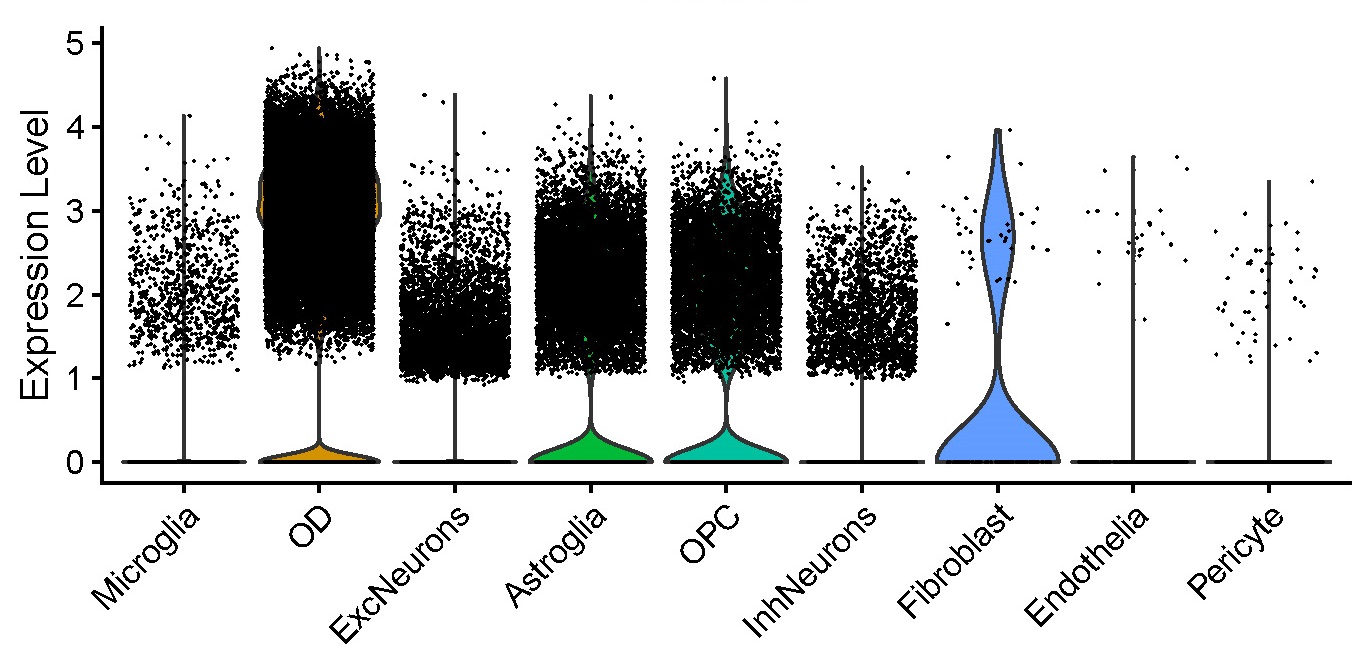

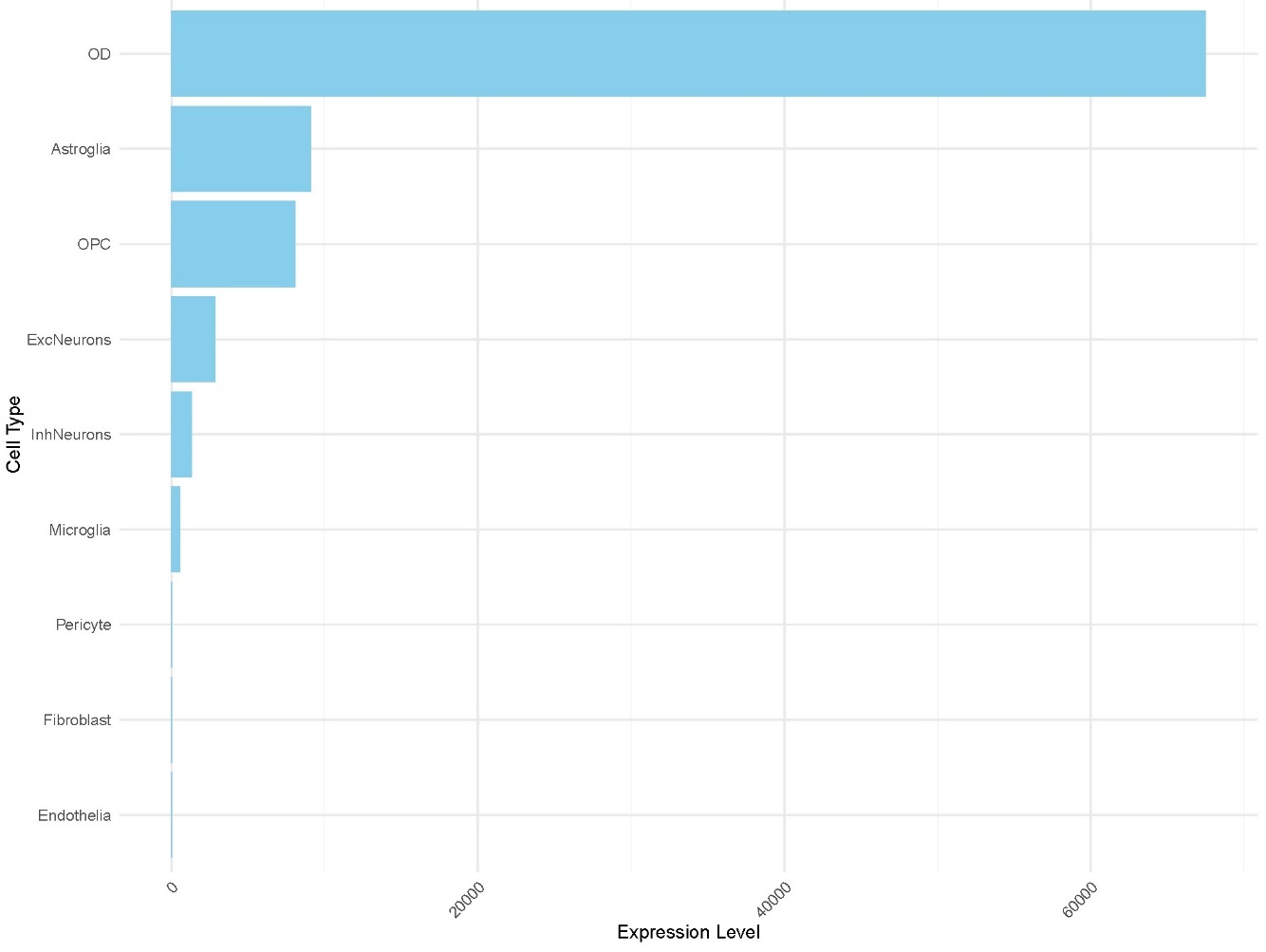

Overall single nucleus RNA expression of *PHLPP1* gene across 9 cell types in human. Oligodendrocytes have the highest expression of *PHLPP1* gene. These plots were created by using a publicly available dataset^4^ (GEO Accession Number: GSE157827).

Abbreviations: OD = oligodendrocytes; ExcNeurons = excitatory neurons; OPC = oligodendrocyte progenitor cells; InhNeurons = inhibitory neurons.

### **Supplementary Figure 6.** Comparison of RNA Expression in OD Between AD Cases and Controls in Humans

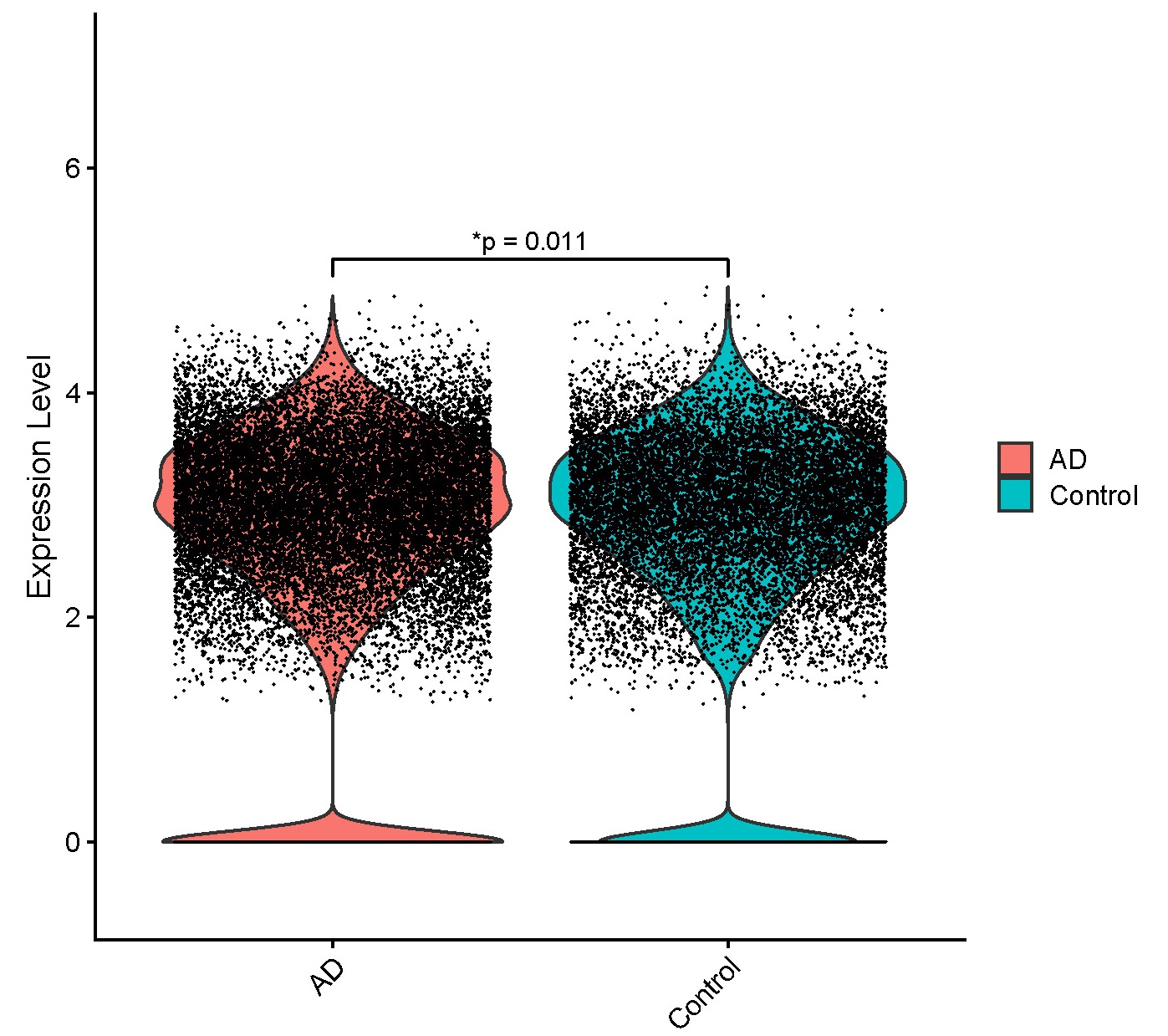

RNA expression of *PHLPP1* gene shows statistically significant changes between AD and Control groups in OD using Wilcoxon rank-sum test.

### **Supplementary Figure 7.** Overall RNA expression of *PHLPP1* across 13 cell types in zebrafish that have been enriched for gliovascular cells

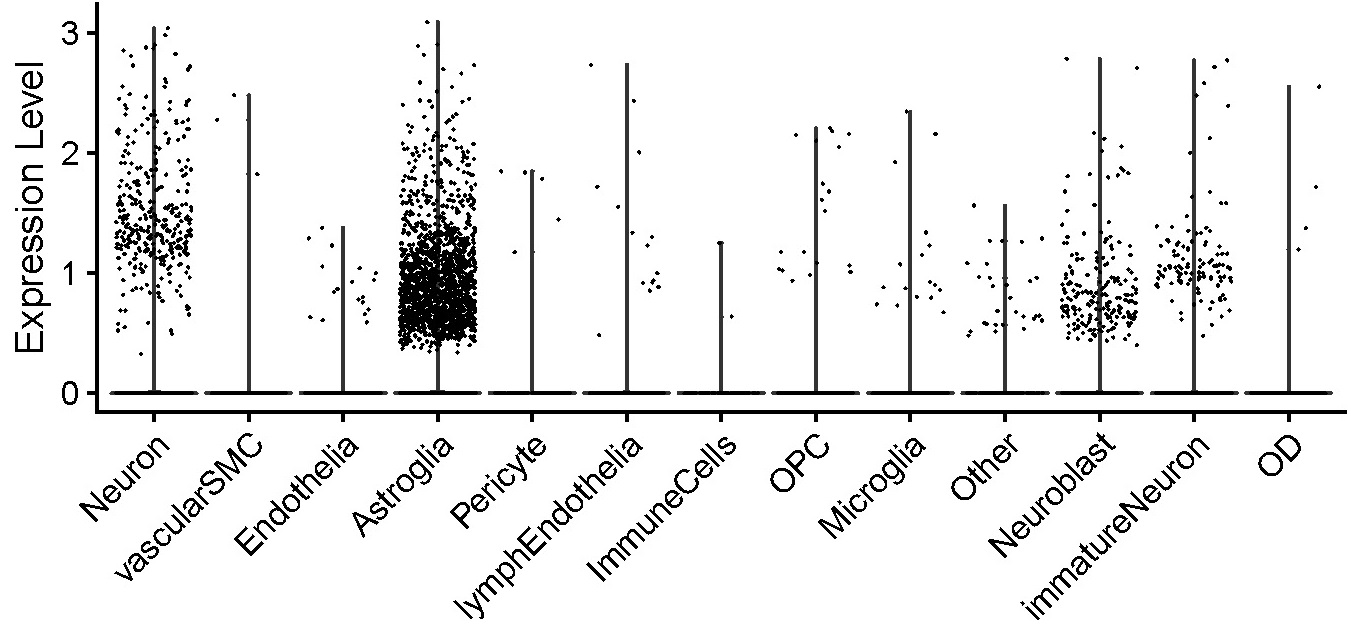

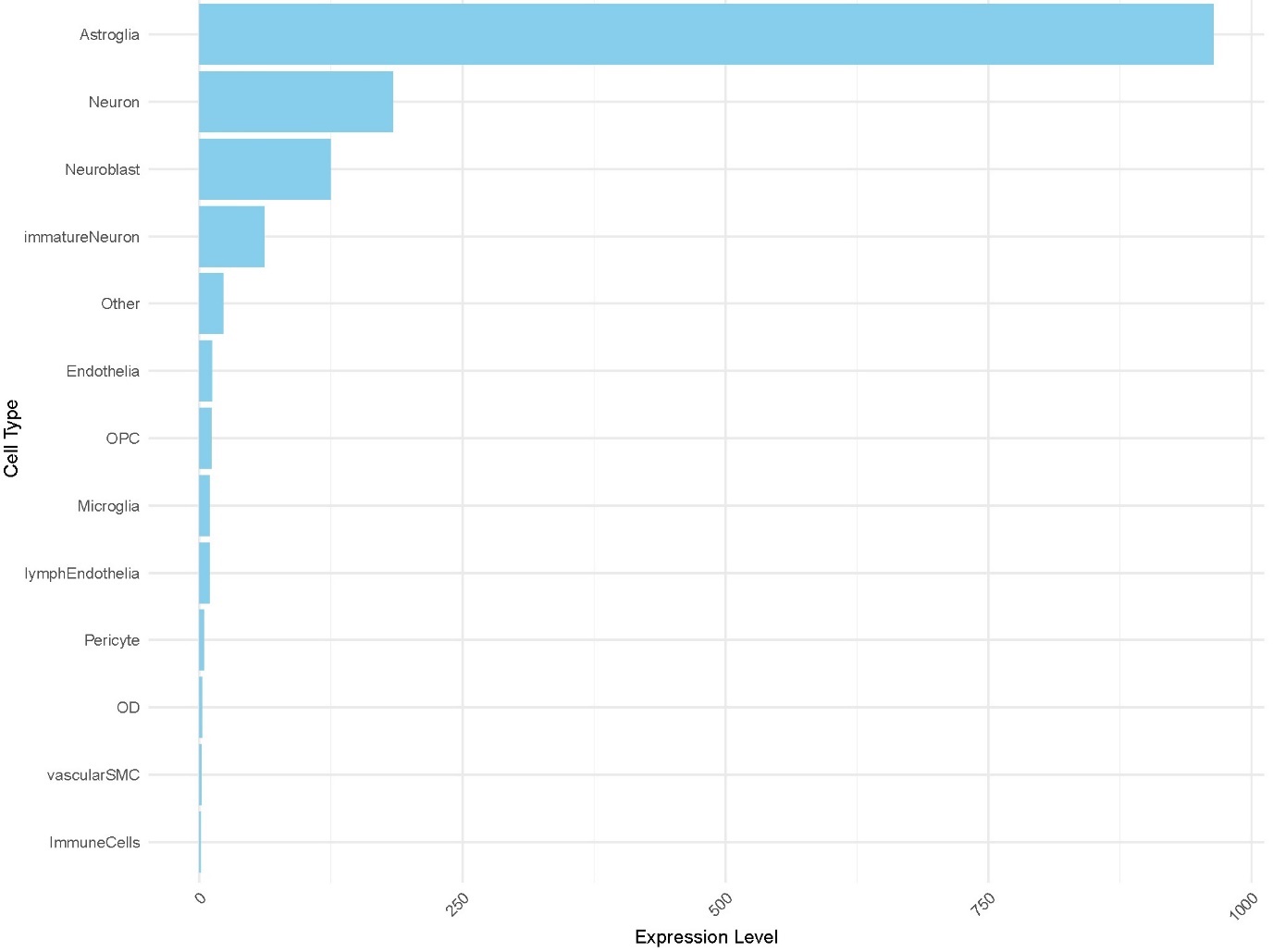

Overall single cell RNA expression of *phlpp1* gene across 13 cell types in zebrafish. Astroglia has the highest expression of *phlpp1* gene in the dataset enriched with gliovascular cell types. These plots have been created by using publicly available dataset^5^ (GEO Accession Number: GSE225721).

Abbreviations: OD = oligodendrocytes; OPC = oligodendrocyte progenitor cells; vascularSMC = vascular smooth muscle cells.

### **Supplementary Figure 8.** Comparison of RNA Expression of *PHLPP1* in Astroglia Between Zebrafish Injected with Aβ42 and Those Injected with Phosphate Buffered Saline (PBS)

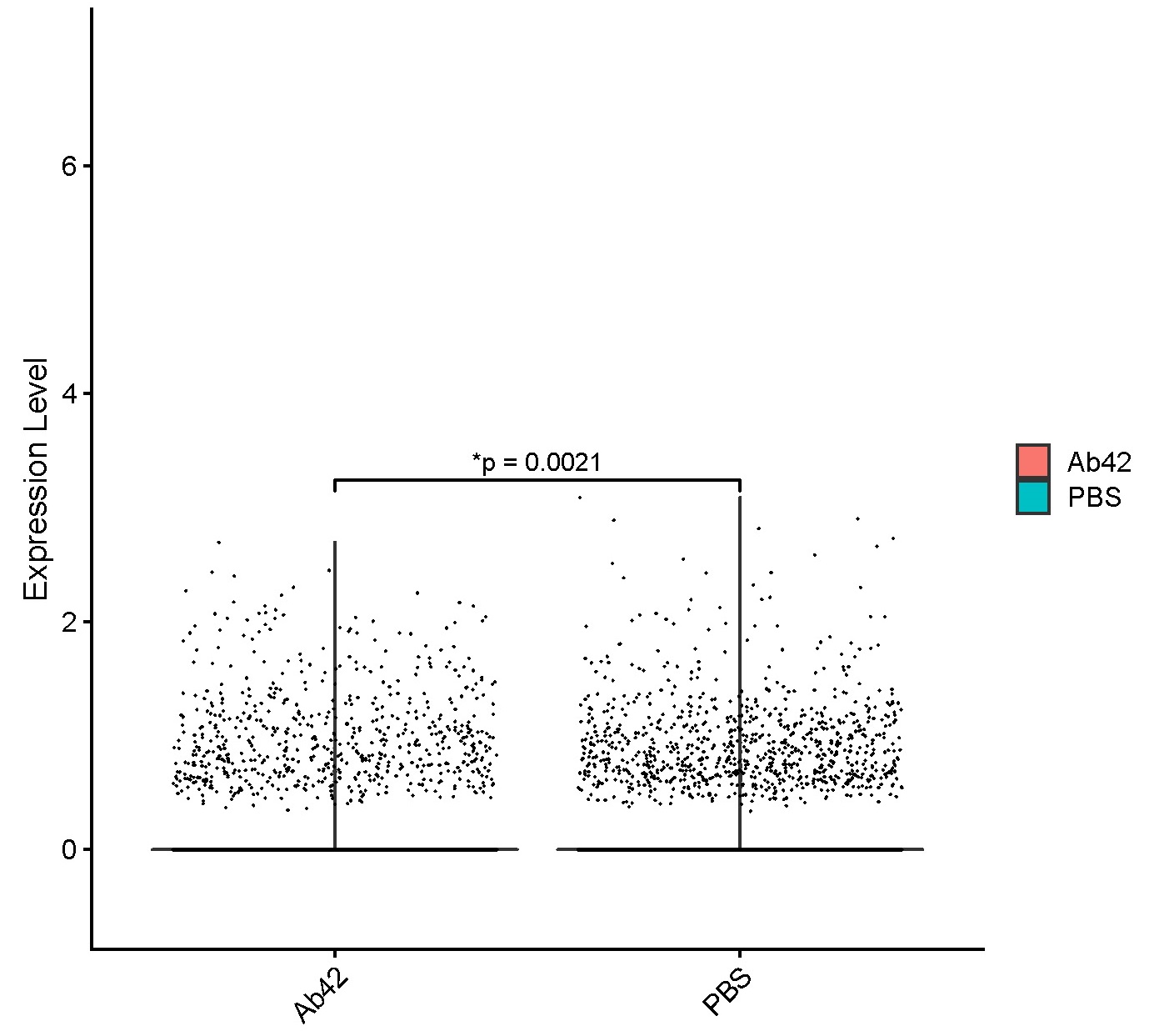

RNA expression of *phlpp1* gene shows statistically significant differences between acute AD model (Ab42 injected) and control (PBS injected) groups in Astroglia using a Wilcoxon rank-sum test.
